# Portuguese Inguinal Hernia Cohort (PINE) study

**DOI:** 10.1101/2020.12.19.20247585

**Authors:** PT Surg – Portuguese Surgical Research Collaborative, J Simões, AA João, JM Azevedo, M Peyroteo, M Cunha, B Vieira, N Gonçalves, J Costa, AS Soares, JS Pimenta, M Romano, AM Cinza, I Miguel, AR Martins, G Fialho, M Reia, FC Borges, CF Monteiro, AC Soares, P Sousa, S Frade, L Matos, JM Carvas, SF Martins, X Sousa, C Rodrigues, JR Carvalho, IC Gil, L Castro, N Rombo, AC Quintela, HM Ribeiro, R Parreira, P Santos, F Caires, A Torre, SC Rodrigues, AH Guimarães, MF Carvalho, MA Pimentel, DC Santos, CF Ramos, C Cunha, C. Azevedo

## Abstract

**Purpose:** Recent comprehensive guidelines have been published on the management of inguinal hernia. Contrary to other European countries, no Portuguese hernia registry exists. This represents an opportunity to assess outcomes of hernia surgery in the Portuguese population. The primary aim is to define the prevalence of chronic pain after elective inguinal hernia repair. The secondary aims are to identify risk factors for chronic pain after elective inguinal hernia repair, to characterise the management of elective inguinal hernia in public Portuguese hospitals.

**Methods:** Prospective national cohort study of patients submitted to elective inguinal hernia repair. The primary outcome is the prevalence of chronic postoperative inguinal pain, according to the EuraHS QoL questionnaire at 3 months postoperatively. The study will be delivered in all Portuguese regions through a collaborative research network. Four 2-week inclusion periods will be open for recruitment. A site-specific questionnaire will capture procedure volume and logistical facilities for hernia surgery.

**Conclusion:** This protocol describes the methodology of a prospective cohort study on the elective management of inguinal hernia. It discusses key challenges and describes how the results will impact future investigation. The study will be conducted across a nationwide collaborative research network, with prospective quality assurance and data validation strategies. It will provide the basis for a more accurate prediction of chronic postoperative inguinal pain and the research on adequate patient selection strategies for surgery and therapeutic strategies for postoperative pain.

## Introduction

The lifetime risk of developing an inguinal hernia is 3-6% for women and 27-43% for men^1^. Symptomatic patients usually undergo surgery, as well as most patients with minimal or no symptoms (approximately 70% ultimately undergo surgery at 5 years^2^). Therefore, surgical repair of inguinal hernia is one of the most common procedures performed by general surgeons worldwide. The development of chronic pain after inguinal hernia repair can lead to severe impairment of patient quality of life and may require re-intervention in a minority of cases. The mechanisms of chronic pain and risk factors are similar for all groin hernias and are often analysed together^3^. PINE will focus in groin hernia including both inguinal and femoral hernias. Chronic pain is commonly defined as pain lasting more than 3 months after hernia repair^4,5^ and is estimated to occur in 10-15% of patients after Lichtenstein hernia repair ^6,7^. The prevalence of chronic pain after inguinal hernia repair in Portugal is unknown.

According to a survey conducted between 2001 and 2005, more than 9,000 surgical procedures for inguinal hernia were performed in Portuguese Public Hospitals^8^. Since then, there has not been an update on the management of inguinal hernia in Portugal. At present, there is no benchmark for the delivery of inguinal hernia surgery on a national level and there is no national hernia registry, as is the case in other countries^9,10^.

Given the high volume of inguinal hernia surgery, it is clinically relevant to define the current approach to inguinal hernia management in Portugal. The characterization of clinical practice variation could highlight the best management processes and allow their dissemination at a national level. From a Health System point of view, this could lead to important savings^11^ and care optimisation.

The recent publication of comprehensive guidelines on the management of inguinal hernia^6^, as well as the existence of national recommendations from the health governmental regulatory body “*Direção Geral de Saúde”* on antibiotic prophylaxis in the context of inguinal hernia repair, offers us an opportunity to query the current practice in Portugal. We believe that this framework could provide actionable insight into such a fundamental area in general surgery.

The PINE study will be conducted by a trainee-led national network of surgical residents. The model for trainee-led research collaboratives was pioneered in the UK. ^12,13^. These networks have proved successful in delivering major surgical research initiatives, including multicentre cohort studies and randomised controlled trials (RCTs)^14^. The Portuguese Surgical Research Collaborative (PT Surg) is a trainee led non-profit organisation, that aims to develop research in surgery, delivered by surgery residents and medical students. It was created to help a new generation of research active, nationally-linked surgeons, setting up a platform for collaboration both at national and international levels (https://twitter.com/pt_surg & www.ptsurg.org). PT Surg has already actively participated in other international studies^15,16^.

A national prospective multicentre cohort study will be conducted to define the risk factors for chronic pain after elective inguinal hernia repair in Portuguese hospitals and will identify current management options.

## Aims

### Primary objective

to define the prevalence of chronic pain after inguinal hernia surgery.

### Secondary objectives

to identify risk factors for chronic pain after elective inguinal hernia repair, to characterise the management of elective inguinal hernia in public Portuguese hospitals.

## Methods

### Study Design

Prospective multicentre cohort study. Patient data will be collected at index admission, 30 days, 3 and 6 months after surgery (*see fig. 1*). This protocol follows the STROBE guidelines ^17^ and the STROCCS statement^18^. PINE study represents a phase 2b study of the surgical innovation framework IDEAL^19^. PINE is registered at Clinicaltrials.gov with the reference NCT04328597.

### Patient selection

#### Inclusion criteria

- Consecutive patients over 18 years of age submitted to elective groin hernia repair in Portuguese Hospitals
- Willing and able to consent and comply with the follow up protocol

#### Exclusion criteria

- Urgent/emergent inguinal hernia repair

Patients will be recruited by the clinical care team at each centre and identified through active search of schedules for elective surgeries. Participant centres will include hospitals of all levels of differentiation and means, ranging from district hospitals to tertiary referral centres. Prior to patient’s enrolment, each centre will complete a survey describing the logistic conditions available, as well as some variables related to surgery volume (see supplementary table 1).

### Patient assessment and outcomes

A member at each study centre will approach the patient during the index admission to explain the objectives of the study and the planned assessment schedule (*see figure 1*). At each centre, in order to obtain ethical approval, additional items may be added to the protocol after revision by the committee. Informed consent will be obtained from all participants. It will be highlighted to the participant that informed consent can be withdrawn at any moment, as well as that there is no obligation to enter the study. The absence of any clinical consequence in case of withdrawal of the informed consent or refuse to participate in the study will also be highlighted to the patient.

At hospital admission, data will be collected on the patient’s demographics, preoperative assessment (*see supplementary table 1*) and surgical approach (*see supplementary table 2*). Patients will be contacted by telephone to gather data on early postoperative period at 30 (±2) days after the index surgery *(see supplementary table 3)*, if the first outpatient clinic appointment falls outside of this period. Data on the late postoperative period at 3 months (±1 week), 6 months (±1 week) after the index surgery (*see supplementary table 4*) will also be assessed in the same way.

Patients will be considered lost to follow up for defined time periods if the study team cannot contact them after 3 attempts in 3 different days.

### Data management and quality assurance

A team of collaborators will recruit patients in each specific period. Data will be collected and stored online through a secure server running the Research Electronic Data Capture (REDCap) web application. REDCap allows collaborators to enter and store data in a secure system. It is widely used by academic institutions throughout Europe and all storage of web-based information by this system is encrypted and compliant with HIPAA-Security Guidelines in the United States. One REDCap login will be issued per recruitment team.

All anonymous data will be held for a total of three years after study completion, after which it will be permanently removed from the server space. A unique ‘REDCap ID’ will be generated by the system for each patient. A local cross-reference of hospital numbers and REDCap IDs should be kept in a secure, encrypted spreadsheet on an institutional password-protected computer. After obtaining the written informed consent from the patient, a contact phone number will be kept for further follow up according to the assessment plan (*see figure 1*). Paper data registries should be destroyed as confidential waste within the center, once uploaded to REDCap. This document should be deleted at the end of the follow-up period.

The local lead is responsible for securing the Ethics Committee approval at each hospital. Only hospitals with confirmed ethical approval will recruit patients.

### Data collection tools

Before starting patient recruitment, centre specific variables will be collected through a centre survey (*see supplementary table 5*).

Baseline and operative patient data will be collected through patient interview and admission record consultation. Inguinal hernias will be classified according to the European Hernia Classification^20^.

After discharge, follow-up will be conducted through phone interviews at 1, 3- and 6- months post-surgery, independently of planned clinic visits (except for the 1^st^ month, as previously explained).

Operative morbidity will be assessed according to the Clavien Dindo classification^21^. Surgical site infection will be defined according to the CDC criteria ^22^ and additionally cases with antibiotic use for surgical site inflammatory signs with no overt pus drainage. If the patient is not able to confirm the occurrence of surgical site infection by phone, medical records and prescriptions will be retrieved from the individual health data platform (*Plataforma de Dados de Saúde*).

Chronic postoperative inguinal pain (CPIP) will be characterized from Patient Reported Outcomes (PRO) data. For this purpose, we will use the EuraHS-QoL score, Portuguese version, developed by the Working Group of the European Registry for Abdominal Wall Hernias (EuraHS)^23^. The EuraHS-QoL questionnaire measures quality of life in patients undergoing abdominal wall hernia repair, with or without mesh implantation to repair the defect. It is employed both pre- and post-operatively. This PRO score is based on a Numerical Rating Scale for three dimensions:

1. pain at the site of the hernia or the hernia repair
2. restriction of activities
3. cosmetic discomfort

Patients will be asked to answer 9 questions comprising the dimensions above. As this questionnaire assesses other quality of life dimensions besides pain, we will be able to describe their variation over time. The score has been validated and EuraHS QoL pain domain was found to be strongly correlated to Visual Analogue Scale and Verbal Rating Scale^23^. The cutoff of 3/10 is the most used to define moderate/disturbing pain^24–28^ and is related to treatment request in such numeric rating scales^29^. Chronic postoperative inguinal pain will be defined as a score of ≥ 3/10 on any of the question of the pain domain in the scale (during rest, during activity, in the last week) 3 months or more after surgery. Patient compliance in answering EuraHS-QoL will be assessed in our Patient and Public Involvement (PPI) exercise, including patients’ ability to answer by phone.

### Data validation

Following data collection, only data sets with >90% data completeness will be accepted for pooled analysis. Centres with >10% missing data points will not be included in the study or in scientific publications referring to this study. A validator will independently identify all patients eligible for inclusion over a 14-day study period. The target for case ascertainment is > 85% and certain key variables will be confirmed.

### Statistical considerations

We anticipate that over 25 centres will participate, each enrolling a minimum of 10 patients during the study period. A cohort of 500 patients would allow to estimate the prevalence of chronic pain after inguinal hernia management (assuming a prevalence of 15.2%)^7^, with 80% power, a significance level of 5% (α=0.05) and a margin of error of ±3%.

The outcomes in relation to our primary objective (chronic pain after elective inguinal hernia repair) will be characterized through descriptive statistics. The outcomes in relation to our secondary objective (characterization of the elective management of groin hernia) will be analysed with descriptive statistics. Logistic regression will be applied to the data to identify risk factors for chronic pain after elective groin hernia repair. The following clinical factors were identified as potential risk factors:

- Age
- Gender
- Pre-operative inguinal pain
- Chronic pain from other cause
- Previous inguinal hernia repair
- Operative technique
- Mesh fixation method
- Nerve handling
- Peri-operative field block
- Immediate post-operative pain
- Post-operative complications (*Clavien Dindo* Classification)
- Inguinal sensitive symptoms

These factors will be studied prior to the logistic regression to identify its association with the presence of chronic pain after elective inguinal hernia repair through bivariate statistical analysis.

An interim analysis will be carried out after the 30-days follow-up assessment is completed for every patient included. No hypothesis testing will occur at this point. Patients lost to follow up, i.e. with missing data on the 3 months postoperative assessment, will not be included in the main analysis.

### PINE study organisational structure

The study will be coordinated by PT Surg, which is the study promotor in collaboration with the Laboratory of Clinical Pharmacology and Therapeutics of the Lisbon School of Medicine. All Portuguese hospitals performing elective inguinal hernia repairs will be invited to participate. A core group of trainees will manage the study. At each centre, there will be a local lead, a supervising consultant and one team in each recruitment period. Each of these teams will have 3 elements, either trainees and/or medical students. Because we aim to promote the development of a national network for conducting clinical research in general surgery, our plan is to organise a course on research methodology to stimulate the scientific development of all the collaborators, therefore adding a benefit for participating in the study. A national and international representative expert advisory panel of surgeons dedicated to abdominal wall surgery has been consulted for the purpose of the developing this protocol and interpreting the findings.

### Patient and public involvement

A selected group of patients has critically reviewed the data collection variables, schedule feasibility and outcomes. The relevance of the outcomes and feasibility of the proposed assessment schedule was discussed and deemed relevant and acceptable by the patients. The patients also agreed that there should be an option to collect their phone numbers. These results will be detailed in a posterior publication.

### Authorship policy

Data collection team collaborators, consultant surgeons, data validators, local leads and the study management group are eligible for PubMed-citable co-authorship^30^. A maximum of three collaborators per data collection period will be listed as ‘PubMed’ citable authors. Validators at each site are also eligible for authorship. Centres with >5% missing data will be excluded from the analysis and the contributing local team will be not be eligible for authorship. In line with the International Committee of Medical Journal Editors authorship guidelines, one consultant per centre is eligible for collaborative PubMed citable co-authorship providing that the following criteria are met: support of local study registration, circulate information about the study to consultant colleagues, and facilitate the presentation of local results at a departmental meeting. For an example of this authorship style, see PubMed with PMID: <u>29897171</u>

### Study schedule

Based on previous projects developed with similar methodology, the planned schedule for the PINE study can be found on table 1.

## Conclusions

This protocol describes the methodology of a prospective cohort study on the elective management of inguinal hernia in Portuguese hospitals. It discusses key challenges and describes how the results will impact future investigation. The study will be conducted across a nationwide collaborative research network, with prospective quality assurance and data validation strategies.

It will provide the basis for a more accurate prediction of chronic postoperative inguinal pain as predictive factors for this postoperative adverse event will be studied. The study will also contribute to the research on adequate patient selection strategies for surgery and therapeutic strategies for postoperative pain.

## Supporting information

Authoship list

## Data Availability

All data is anonymized and secured in an on-line database (Redcap)

## Acknowledgments

We thank our expert advisory panel (A de Beaux, E Guerreiro, F Ferreira, F Muysoms, H Friis-Andersen, M Rosen, N Henriksen, S Morales-Conde, T Bisgaard) for critical review of the protocol and constructive feedback.

We thank T Pinkney for helpful discussion on the methodology of this project.

We thank our patient advisory panel for critical review of the data collection variables, schedule feasibility and outcomes.

We thank our expert advisory panel (A de Beaux, E Guerreiro, F Ferreira, H Friis-Andersen, M Rosen, N Henriksen, S Morales-Conde, T Bisgaard) for critical review of the protocol and constructive feedback. We thank our patient advisory panel (A Rocha, J Fonseca) for critical review of the data collection variables, schedule feasibility and outcomes.

## Supplementary documents

**Supplementary table 1.** Demographic data and Surgical assessment.

| Variable name |  | Options | Description |
| --- | --- | --- | --- |
| Demographic data | Age |  | In years |
|  | Gender |  | Male, Female |
|  | BMI |  | Weight / Height squared |
|  | ASA grade | I, II, III, IV |  |
|  | Smoking habits | Yes, No, Ceased |  |
|  | Number of pack years |  | (packs smoked per day) × (years as a smoker) |
|  | Family history of inguinal hernia | Yes, No |  |
|  | Comorbidities | Abnormal collagen metabolism |  |
|  |  | Immunodepression |  |
|  |  | Pulmonary comorbidities | COPD, asthma |
|  |  | Cardiovascular comorbidities | Ischemic heart disease, valvulopathy, congestive heart failure |
|  |  | Hepatic disease | Cirrhosis, ascites |
|  | Chronic pain from other cause (not the hernia) | Headache, Inflammatory osteoarticular disease, Degenerative osteoarticular disease, fibromyalgia, post-traumatic pain, post-surgical pain, cancer, nervous injury/compression, other |  |
|  | Previous inguinal hernia repair | Yes/No |  |
|  | Number of previous surgeries |  |  |
|  | Site of previous repair | Ipsilateral, Contralateral |  |
|  | Approach | Anterior, posterior, laparo-endoscopic |  |
|  | Time since last surgery |  | In months |
|  | Previous prostatectomy | Yes, No |  |
| Surgical assessment | Indication for surgery | Asymptomatic or Minimally symptomatic; Symptomatic | Symptomatic defined as painful, emergency department consultation, interference with daily life |
|  | Preoperative imaging | Yes/No |  |
|  | Type of imaging | Ultrasound, CT, MRI, herniography |  |

**Supplementary table 2.**
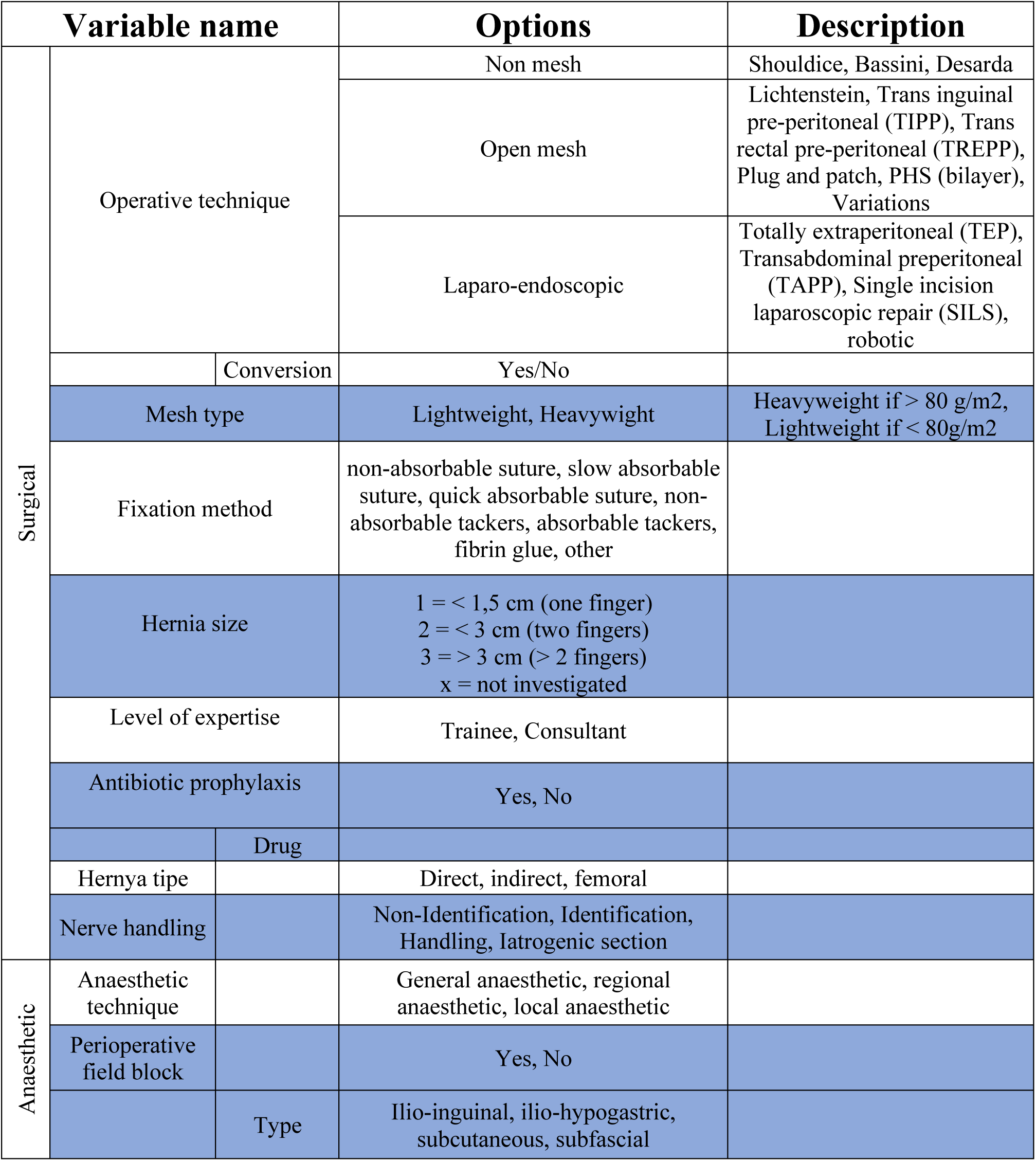
Operative data variables.

**Supplementary table 3.** Early (30 days) postoperative assessment data.

| Variable name |  | Options | Description |
| --- | --- | --- | --- |
| Discharge dispositions | Discharge after surgery | Same day, next day, other | If other, specify number of days |
|  | Immediate post operative pain | numerical grading scale (0 - 10) | (maximum level during first week) |
|  | Post discharge analgesia | Opioid, NSAID, Other | specify other |
|  | Time to return to work |  | Time to return to work (days) |
|  | If hasn't returned specify reason |  |  |
| Pain | Inguinal sensitive symptoms | Parestesia, Anesthesia | (in the operated inguinal region) |
|  | EuraHs QoL Score [22] |  | http://eurahs.eu/EuraHS-QoL-download.php |
| Complications | Surgical site infection [21] | Yes, No | → Purulent with or without laboratory confirmation, from the superficial incision<br>→ Organisms isolated from an aseptically obtained culture of fluid or tissue from the superficial incision<br>→ ≥1 signs/symptoms of infection: pain, localized swelling, redness, or heat<br>→ Diagnosis of SSI by the surgeon |
|  | Return to emergency department | Yes, No |  |
|  | Hospital readmission | Yes, No |  |
|  | Clavien dindo grade | I, II, III, IV |  |

**Supplementary table 4.** 3- and 6-month postoperative assessment data.

| Variable name | Options | Description |
| --- | --- | --- |
| Contact date |  | Date |
| EuraHs QoL Score [22] |  | http://eurahs.eu/EuraHS-QoL-download.php |
| Return to work after surgery | Yes, No |  |
| Time |  | Time to return to work (days) |
| Recurrence | Yes, No |  |

**Supplementary table 5.**
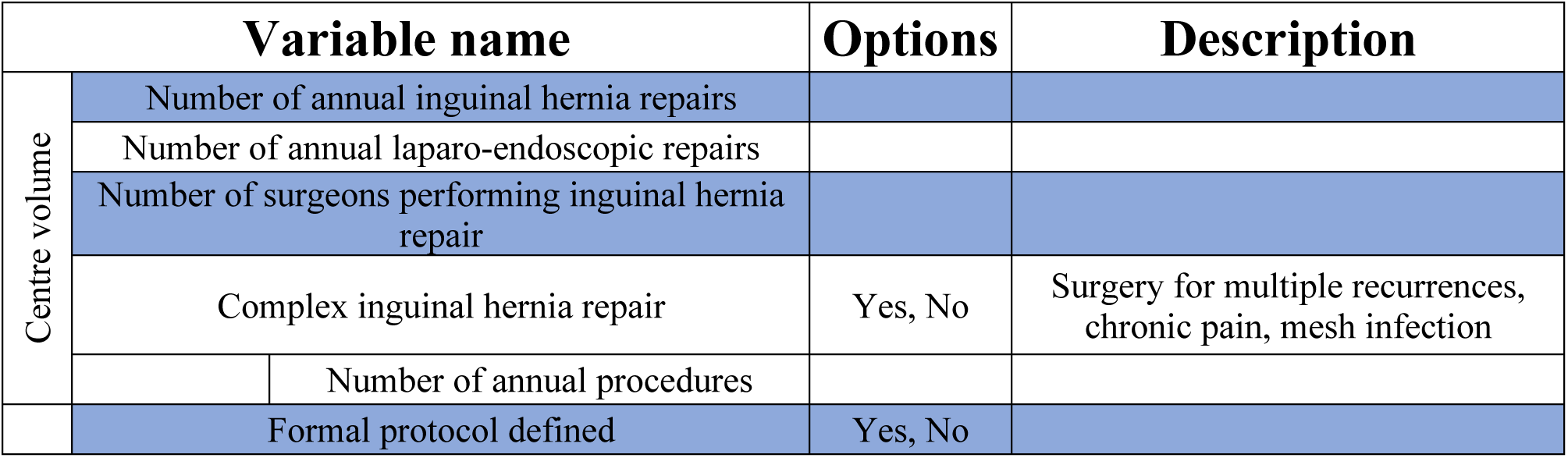
Centre survey.

| Variable name |  | Options | Description |
| --- | --- | --- | --- |
| Centre volume | Number of annual inguinal hernia repairs |  |  |
|  | Number of annual laparo-endoscopic repairs |  |  |
|  | Number of surgeons performing inguinal hernia repair |  |  |
|  | Complex inguinal hernia repair | Yes, No | Surgery for multiple recurrences, chronic pain, mesh infection |
|  | Number of annual procedures |  |  |
|  | Formal protocol defined | Yes, No |  |

**Supplementary table 6.** PINE study schedule.

| Task | Duration | Predicted conclusion date |
| --- | --- | --- |
| Centre recruitment | 3 months | 4th October 2019 |
| Local ethical approvals | 3 months | 4th October 2019 |
| Patient recruitment | 3 periods |  |
| Period 1 | 2 weeks | 7-18 Oct 2019 |
| period 2 | 2 weeks | 28th Oct – 8th Nov 2019 |
| period 3 | 2 weeks | 18 - 29th Nov 2019 |
| period 4 | 2 weeks | 2 - 14th Dec 2019 |
| Interim analysis | 1 month | After 29th Dec 2019 |
| Final analysis | 6 months | After 29th May 20 |

**Suplementary Figure 1.**
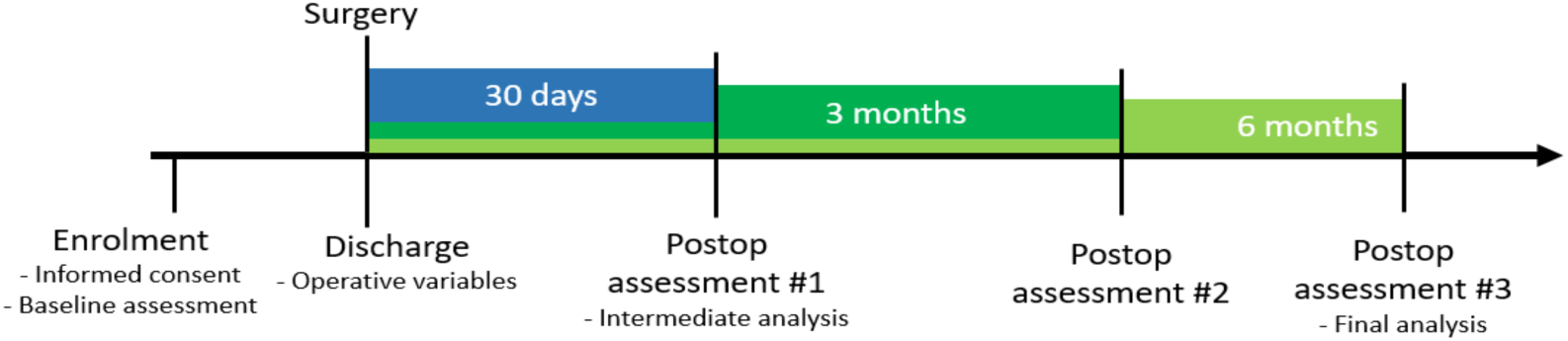
Patient assessment schedule.

