## Supplementary material for "Portuguese Inguinal Hernia Cohort (PINE) study": Authoship list

### Authorship

Steering Committee:

J Simões^1,2^, AA João^1,3^, JM Azevedo^1,4^, M Peyroteo^1,5^, M Cunha^1,6^, B Vieira^1,7^, N Gonçalves^1,8^, J Costa^1,8^, AS Soares^1,3^ (corresponding author, +351963864555)

Local Leads:

JS Pimenta^9^, M Romano^10^, AM Cinza^11^, I Miguel^6^, AR Martins^3^, G Fialho^12^, M Reia^13^, FC Borges^2^, CF Monteiro^14^, AC Soares^7^, P Sousa^15^, S Frade^16^, L Matos^17^, JM Carvas^18^, SF Martins^19^, X Sousa^20^, C Rodrigues^4^, JR Carvalho^21^, IC Gil^21^, L Castro^22^, N Rombo^23^, AC Quintela^24^, HM Ribeiro^25^, R Parreira^26^, P Santos^27^, F Caires^28^, A Torre^29^, SC Rodrigues^30^, AH Guimarães^31^, MF Carvalho^32^, MA Pimentel^33^, DC Santos^34^, CF Ramos^35^, C Cunha^36^, C. Azevedo^37^

Affiliations:

1- PT Surg – Portuguese Surgical Research Collaborative
2- Hospital Garcia de Horta
3- Hospital Professor Doutor Fernando Fonseca
4- Hospital da Horta
5- Instituto Português de Oncologia - Porto
6- Centro Hospitalar Universitário do Algarve - Portimão
7- Hospital do Santo Espírito da Ilha Terceira
8- Faculdade de Medicina da Universidade de Lisboa, Laboratório de Farmacologia Clínica e Terapêutica.
9- Unidade Local de Saúde Baixo Alentejo
10- Unidade Local de Saúde Castelo Branco
11- Hospital do Espirito Santo
12- Hospital de Portalegre
13- Hospital de Elvas
14- Unidade Local de Saúde do Alto Minho
15- Hospital Particular do Algarve
16- Centro Hospitalar Universitário de Lisboa Central
17- Centro Hospitalar de Entre o Douro e Vouga
18- Unidade Local de Saúde do Nordeste
19- Hospital Distrital de Santarém
20- Centro Hospitalar de Setúbal
21- Centro Hospitalar de Lisboa Ocidental - Hospital São Francisco Xavier
22- Centro Hospitalar de Lisboa Ocidental - Hospital Egas-Moniz
23- Centro Hospitalar de Lisboa Ocidental - Hospital de Santa Cruz
24- Hospital Pedro Hispano
25- Hospital Distrital da Figueira da Foz
26- Hospital do Divino Espirito Santo - Ponta Delgada
27- Centro Hospitalar do Oeste - Hospital de Torres Vedras
28- Centro Hospitalar do Oeste - Hospital das Caldas da Rainha
29- Hospital de Gaia
30- Hospital de São João
31- Centro Hospitalar Universitário de Coimbra
32- Hospital de Famalicão
33- Hospital de Aveiro
34- Unidade Local de Saúde do Litoral Alentejano
35- Centro Hospitalar de Lisboa Norte
36- Hospital Beatriz Ângelo
37- Hospital Cova da Beira
